# Association between Depression and Quality of Life among aging population in Bharatpur Metropolitan City, Nepal

**DOI:** 10.1101/2025.10.29.25339108

**Authors:** Nikita Baral, Bipindra Pandey, Samiksha Dhakal, Samarpan Adhikari, Jiwan Kumar Poudyal, Kanchan Thapa

**Affiliations:** Department of Public Health, Shree College of Technology, Bharatpur, Chitwan, Nepal; Deparment of Public Health, Chitwan Medical College, Bharatpur, Chitwan, Nepal; Deparment of Pharmacy, Madan Bhandari Academy of Health sciences, Hetauda, Nepal; Depatment of Chemistry, Tribhuwan University, Bharatpur, Chitwan, Nepal; Public Health Officer, District Public Health Office, Makwanpur,Nepal; Noble Shivapuri Research Institute, Kathamandu, Nepal

**Keywords:** Aging Population, Depression, Geriatric, Health, Nepal, Quality of Life

## Abstract

**Background:** Depression and quality of life (QoL) are significant issues affecting the elderly population. This research sought to evaluate the prevalence of depression and explore its connection to QoL among older adults living in Bharatpur Metropolitan City, Nepal.

**Methods:** A community-based cross-sectional study was carried out from February 2024 to July 2025, involving 250 elderly individuals aged 60 and above. The participants were chosen through a multistage sampling technique. Data collection was conducted using a semi-structured questionnaire, the 15-item Geriatric Depression Scale (GDS-15), and the WHO Quality of Life-BREF (WHOQOL-BREF) instrument. For data analysis, a variety of methods were employed, including descriptive statistics, chi-square tests, ANOVA, Mann-Whitney U tests, correlation analyses, and binary logistic regression.

**Results:** The prevalence of depression among elderly individuals was found to be 28.8%. Factors such as age, gender, marital status, living conditions, level of education, income source, job, chronic illnesses, and alcohol use were significantly linked to depression (p<0.05). Psychological and social life aspects were closely associated with enhanced quality of life (QoL) (p<0.0001 and p=0.018, respectively). Depression had a notable impact on all QoL dimensions, with the environmental aspect being the most affected (OR=17.37, 95% CI: 7.99-43.70).

**Conclusion:** Depression is prevalent among the elderly in Bharatpur Metropolitan City, significantly impacting multiple aspects of their quality of life (QoL). Psychological and social elements are crucial in shaping the QoL of older individuals. To address geriatric depression and improve the overall QoL of Nepal’s aging population, it is recommended to implement screening programs, establish social support systems, and offer training for primary healthcare professionals.

## Introduction

In 2019, there were one billion people worldwide who were 60 years old or older. This number is expected to grow to 1.4 billion by 2030 and further increase to 2.1 billion by 2050.(1) South Asia is projected to undergo a swift aging process, outpacing the rates seen in the last four to five decades. In Nepal, the Senior Citizen Act of 2006 classifies individuals aged 60 and above as senior citizens. This law aims to offer special care to those who are helpless, need special attention, or are unable to care for themselves, including those requiring day care or additional support.(2)

Based on the 2021 census data from Nepal, individuals aged 60 and above constitute 10.2% of the nation’s population.(3) A study conducted by the United Nations Economic and Social Commission for Asia and the Pacific (UNESCAP) in June 2002 on aging trends in Asia and the Pacific indicated that, although the current proportion of elderly people is around 5–9%, this figure is anticipated to rise considerably over the next few decades, potentially doubling the number of those aged 60 and older within 50 years. According to the 2021 census, Nepal’s elderly population stands at 2.97 million, reflecting a 38.2% increase since the 2011 census.(4)

As the population continues to expand rapidly, older adults are facing a multitude of challenges related to healthcare, social support, mental well-being, and more, which are negatively impacting their Quality of Life (QoL). Furthermore, the increasing incidence of non-communicable diseases (NCDs), overseas employment, financial hardships, insufficient infrastructure, and age-related health issues are further diminishing the QoL among seniors. The quality of life is a critical issue for the elderly population.(5)Their quality of life is influenced by various physical, mental, and social elements. Important factors affecting subjective quality of life include mental health issues, symptoms of anxiety and depression, and social connections.(6) Generally, older individuals perceive their quality of life positively based on social interactions, dependence, health, social circumstances, and comparisons with others. While cultural differences in the subjective aspect of quality of life are minimal, differences have been noted in objective aspects. Depression and dementia are two significant factors that greatly influence the quality of life in older adults.(5)It is crucial to address the healthcare needs and service utilization for the elderly in Nepal.(7)Social connections are found to be lacking among people living with diabetes in Nepal.(8)

Population aging is a well-known phenomenon impacting nations worldwide, irrespective of their level of development. In less developed areas, the proportion of older adults is rising due to changes in demographic trends. Nevertheless, their conditions are deteriorating because of the rapid breakdown of traditional family systems, coupled with fast-paced modernization and urbanization. In Nepal, people aged sixty and above are classified as elderly, as per the Senior Citizens Act, 2063.(9)

As a result of changes in epidemiology, chronic diseases and various lifestyle-related disorders are increasingly taking the place of communicable diseases (CDs). Older adults are especially susceptible to depression, social instability, loneliness, and chronic co-morbid conditions.(10) Among the elderly, depression is the most prevalent emotional disorder, greatly impacting their functional abilities and reducing their quality of life (QoL).(11) In Nepal, many elderly people often find themselves without caregivers, highlighting a significant gap in the support and care available to this age group.(12)

In 2017, research conducted in the Kailali district revealed that 45.9% of older participants described their quality of life as neutral, neither positive nor negative. Meanwhile, 35.1% perceived it as good, and 19.0% regarded it as poor.(13) Furthermore, a 2022 study by Chinese researchers, which analyzed data from multiple countries using depression screening tools, underscored the absence of a thorough systematic review on the prevalence of depression among the elderly. The global prevalence of depression has been documented at 28.7%.(14) Similarly, a 2019 study conducted in Nepal’s Kavrepalanchowk district found that 53.1%(11) of the elderly population suffered from geriatric depression. Nepal is categorized by the World Bank as one of the poorest and most vulnerable countries globally. The WHO Study on Global AGEing and Adult Health (SAGE) reports that 4.7% of older adults in low- and middle-income countries (LMICs) experience depression. In Nepal, however, depression rates among the elderly vary significantly: from 17.3% to 89.1% in care facilities, 25.5% to 60.6% among those living in the community, and 53.2% to 57.1% among older patients in hospitals.(15)

Even with the assistance provided by some non-governmental sectors, older adults encounter a range of physical, mental, and psychological difficulties, such as diabetes, high and low blood pressure, memory decline, depression, and physical impairments. The lack of a caregiver worsens these problems, making the aging process more challenging for seniors and reducing their overall quality of life. The government’s insufficient efforts in creating nursing homes or other programs for the elderly highlight the urgent need to address the complexities and requirements of Nepal’s aging population.(12)

There are limited government and community programs available, including senior care facilities, pensions for the elderly, and financial support for older individuals. However, there is a lack of documentation regarding the challenges in implementing these programs and their overall effectiveness. (16) Consequently, this study seeks to determine how depression affects the overall quality of life of elderly individuals residing in specific wards of a newly developed metropolitan city of Nepal.

## Materials and Methodology Study design and settings

A cross-sectional analytical study based in the community was conducted from February 2024 to July 2025 in selected wards of Bharatpur Metropolitan City, situated in the southern region of Bagmati Province in Chitwan District.(17) This city, which serves as the district headquarters for Chitwan, is noted as Nepal’s second-largest metropolitan area by land size and comprises 29 wards. According to the national census of 2021, Bharatpur Metropolitan City has a total population of 369,268, consisting of 178,897 males (48.4%) and 190,371 females (51.6%), which includes 20,583 elderly females and 19,020 elderly males.(18) Bharatpur Metropolitan City is one of the crowded cities of Nepal. Many people live here. It is one of the most rapidly urbanized cities. There is a high rate of immigration, particularly from the hilly region. Therefore, different characteristics of people come here with their families to make use of the available resources and facilities. Elderly people experience mental health issues, including depression, as a result of their children moving away in search of better career opportunities. Elderly people are left behind because of their inability to understand the fast-paced evolution of technology and lifestyle. They live alone and face various problems such as loneliness, depression, and many other physical activities. Depression affects the quality of life of many elderly people. Depression results in a poor quality of life.

## Study Population

This study was carried out among an elderly population of both sexes aged 60 years and above from Bharatpur Metropolitan City, residing for at least six months in this research methodology. One elderly person was chosen randomly in the case of two or more elderly people in one household. Participants who were deaf and dumped, seriously ill, and unwilling to provide consent were excluded from the study.

### Sample size

The sample size was determined using Cochran’s (19)corrected formula, which is appropriate for finite populations when assessing proportions. Considering that 82.4% (p = 82.4%) of older adults report a fair quality of life,[19] a 5% margin of error and a 95% Confidence Interval (CI) were employed for the study. By including an additional 10% to account for possible non-responses, the final sample size set for this research was 250.

### Sampling technique

This study employed a multistage sampling approach to enlist the participants. Bharatpur Metropolitan City was intentionally chosen. From the twenty-nine wards, four wards (ward numbers 4, 12, 16, and 22) were selected through simple random sampling using the lottery method. The four elected wards (Ward 4, Ward 12, Ward 16, and Ward 22) consisted of a total of 7078 elderly population, and these chosen wards consisted of n_4_ =2403 elderly population, n_12_ = 1802 elderly, n_16_ =2099, and n22= 774 elderly population, respectively. After selecting the wards, the sample was selected based on the total elderly participants of the respective wards. The sample sizes for each ward were 85, 64, 74, and 27, respectively. Purposive sampling was applied to approach the respondents. The same process was repeated in every selected ward.

## Data collection Tools and measures

A semi-structured questionnaire was administered to collect the necessary information. It featured inquiries concerning socio-demographic characteristics. To assess the quality of life (QoL) and depression in older adults, established evaluation tools were employed. The WHOQOL-BREF instrument was used to gauge the quality of life among seniors, while the validated Nepali version of the GDS-15 scale served to identify depressive symptoms in this group. The Quality-of-Life questionnaire was divided into four sections (Annex A). In this research, a translated Nepali version of the survey was utilized (Annex B). Additionally, the variables related to QoL, Depression, Elderly, and Education were systematically organized for analysis (Annex C).

## Data Collection

Interviews conducted face-to-face, each lasting approximately 25 to 30 minutes, were used to gather data. The data collection was managed by NB, a BPH student, and took place between April 1 and July 30, 2024. Structured questionnaires were utilized to evaluate the quality of life and depression among the elderly. The WHOQOL BREF(20)tool was employed to collect data on quality of life, while the validated Nepali version of the GDS-15(21) scale was used to measure depression in the elderly population.

## Data management and analysis

The data collected were evaluated for their accuracy, completeness, and relevance, and were then organized and coded on the day they were gathered to facilitate seamless data entry. Subsequently, the information was input into the Statistical Package for the Social Sciences (SPSS) IBM Version 22. For the purpose of data analysis, both IBM SPSS Version 22 and R Studio (version 4.3) were employed. All the data used in this analysis are publicly available for further review at https://doi.org/10.7910/DVN/JMHORK . Similarly, researcher also adhered on STROBE checklist to report this study findings (Annex E)

The analysis adhered to the GDS-15 and WHOQOL BREF guidelines. The study’s findings are detailed in the results section. Descriptive statistics, such as frequency, percentage, mean, and median, were computed to summarize the socio-demographic and health-related characteristics of the elderly participants. The Chi-square test was used to examine the association between categorical variables like gender, marital status, educational level, and classifications of quality of life as well as depression status.

ANOVA was employed to assess the mean scores of continuous variables across several groups. For comparing two independent groups when the data did not adhere to a normal distribution, the Mann–Whitney U test was used as a non-parametric alternative to the t-test. Furthermore, a correlation test was performed to investigate the relationship between continuous variables, such as the connection between age and WHOQOL-BREF scores or between depression scores (GDS- 15) and quality of life (QoL) scores. This research also applied binary logistic regression to evaluate the influence of depression on different quality of life aspects. All four dimensions were categorized into Good QoL and Bad QoL based on their mean scores, which were further examined for the impact of depression. The findings are presented in a forest plot, along with Odds Ratios (OR) within a 95% Confidence Interval (CI).

## Ethical consideration

The Institutional Review Committee at Shree College of Technology Pvt. Ltd. approved this research [Reference No: SMTC-IRC-20240319-72]. A letter endorsing data collection was obtained from Bharatpur metropolitan city. Before gathering data, participants received a thorough explanation of the study’s aims. Both written and oral consent were secured from each participant. It was made clear to participants that the data collected would be utilized solely for research purposes. The study adhered to social and cultural values, ensuring no discrimination occurred based on caste, religion, socioeconomic status, or any other factors. Participant confidentiality was maintained by keeping their identities anonymous, ensuring their names were not revealed.

## Results

Table 1 shows that the highest proportion (46.0%) of participants were young old (60-69 years), 50.4% were male, and 68.8% were married. The majority of participants (94.4 %) followed Hinduism, 11.6% were Brahmin/Chhetri, and 71.2% resided in joint families. Furthermore, 46.8 elderly had basic and above level education, 39% were illiterate, and 14% had informal education. The main source of income was allowance (51.2 %). The majority of the elderly, 35.6% were not able to work.

**Table 1:**
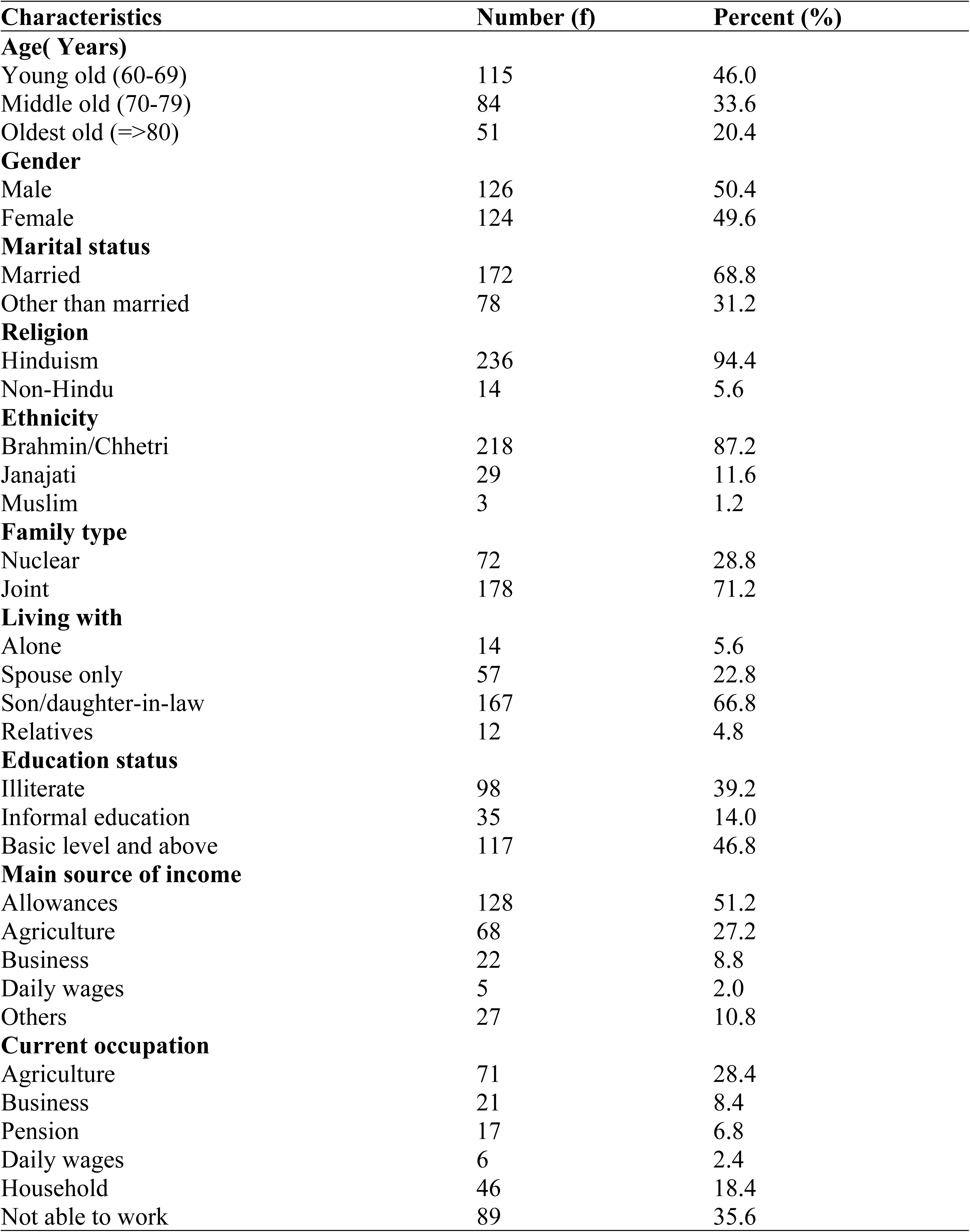
Socio-economic and demographic characteristics of participation (n=250)

| Characteristics | Number (f) | Percent (%) |
| --- | --- | --- |
| **Age( Years)** |  |  |
| Young old (60-69) | 115 | 46.0 |
| Middle old (70-79) | 84 | 33.6 |
| Oldest old (>=80) | 51 | 20.4 |
| **Gender** |  |  |
| Male | 126 | 50.4 |
| Female | 124 | 49.6 |
| **Marital status** |  |  |
| Married | 172 | 68.8 |
| Other than married | 78 | 31.2 |
| **Religion** |  |  |
| Hinduism | 236 | 94.4 |
| Non-Hindu | 14 | 5.6 |
| **Ethnicity** |  |  |
| Brahmin/Chhetri | 218 | 87.2 |
| Janajati | 29 | 11.6 |
| Muslim | 3 | 1.2 |
| **Family type** |  |  |
| Nuclear | 72 | 28.8 |
| Joint | 178 | 71.2 |
| **Living with** |  |  |
| Alone | 14 | 5.6 |
| Spouse only | 57 | 22.8 |
| Son/daughter-in-law | 167 | 66.8 |
| Relatives | 12 | 4.8 |
| **Education status** |  |  |
| Illiterate | 98 | 39.2 |
| Informal education | 35 | 14.0 |
| Basic level and above | 117 | 46.8 |
| **Main source of income** |  |  |
| Allowances | 128 | 51.2 |
| Agriculture | 68 | 27.2 |
| Business | 22 | 8.8 |
| Daily wages | 5 | 2.0 |
| Others | 27 | 10.8 |
| **Current occupation** |  |  |
| Agriculture | 71 | 28.4 |
| Business | 21 | 8.4 |
| Pension | 17 | 6.8 |
| Daily wages | 6 | 2.4 |
| Household | 46 | 18.4 |
| Not able to work | 89 | 35.6 |

Table 2 shows that the majority, 68.4 %, had chronic diseases. Among these participants, 18.4% had a smoking habit and 8% had an alcohol consumption habit. Similarly, 28.8% had a depression status (GDS≥6).

**Table 2:** Distribution of diseases and related behavioral factors of respondents (n=250)

| Characteristics | Number(f) | Percent (%) |
| --- | --- | --- |
| Presence of Chronic disease | 171 | 68.4 |
| Currently smoking | 46 | 18.4 |
| Consume Alcohol | 20 | 8.0 |
| Presence of Geriatric Depression (GDS≥6) | 72 | 28.8 |
| Types of chronic disease (n=171)* |  |  |
| Respiratory related disease | 20 | 11.7 |
| High blood pressure | 111 | 64.9 |
| Heart related disease | 23 | 13.5 |
| Diabetes | 46 | 26.9 |
| Gastrointestinal disease | 3 | 1.8 |
| Thyroid | 21 | 12.3 |
| Others | 3 | 1.8 |

\*Respondents with chronic illness only

Table 3 illustrates the link between socio-demographic factors and geriatric depression, as determined by chi-square and Fisher’s exact tests. In a similar vein, gender was found to have a significant association with depression in the elderly. The study also identified a notable connection between marital status and depression among older adults. Additionally, the living conditions of seniors were meaningfully related to depressive symptoms. The research highlighted a strong relationship between the educational attainment of older individuals and their mental health in terms of depression. Similarly, the type of income was significantly linked to depression in the elderly demographic. The findings further showed that current employment status had a notable association with geriatric depression. Moreover, drinking habits among older adults were significantly connected to their depressive states.

**Table 3:**
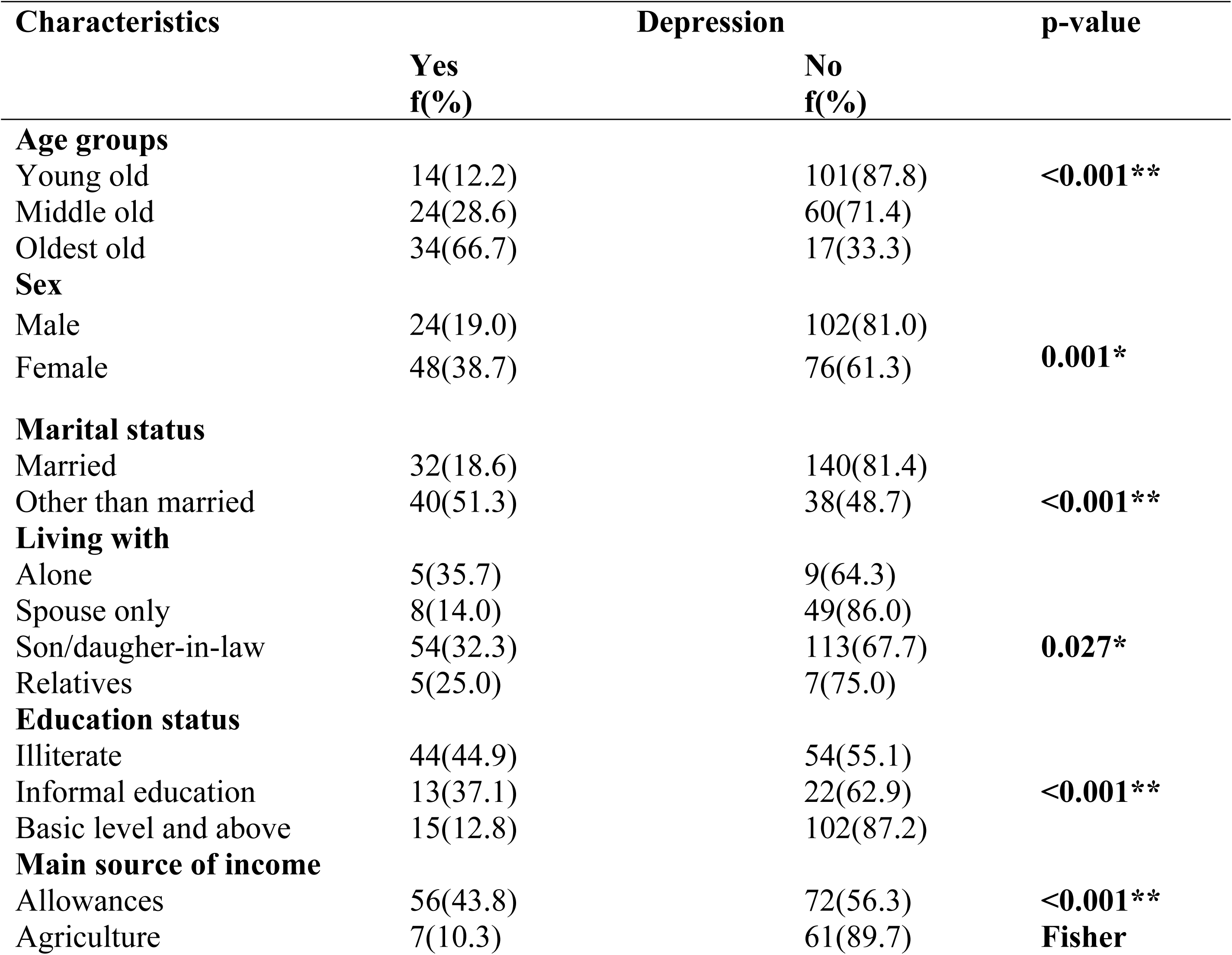
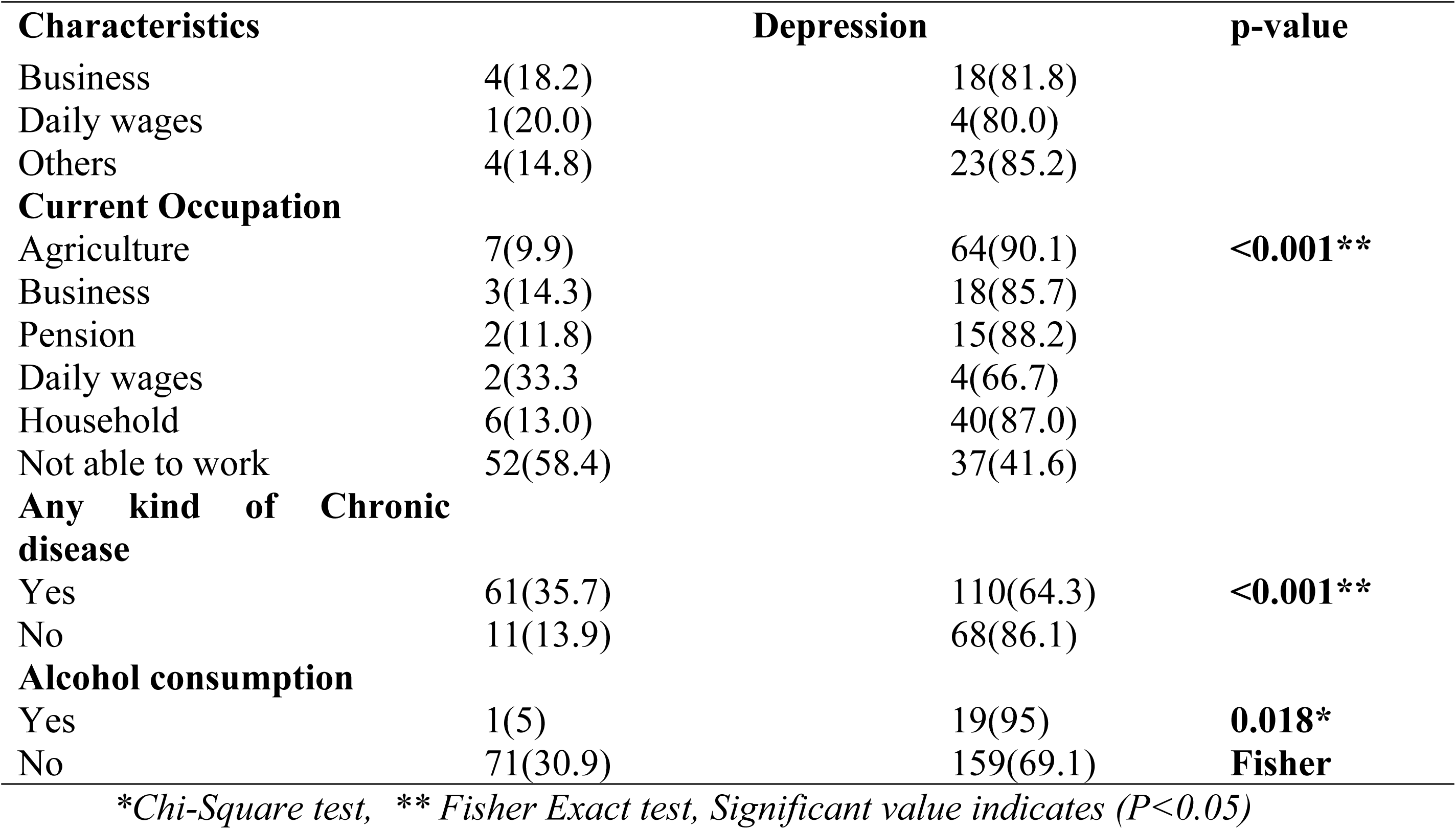
Association between Geriatric depression and Socio-demographic factors (n=250)

Figure 2 illustrates the influence of various health domains—physical, social, psychological, and environmental—on the overall quality of life (QoL) among elderly participants (n = 250). To evaluate the differences in QoL between those categorized as having poor and good quality of life, the Mann–Whitney U test was utilized. Participants with a good QoL exhibited a higher mean rank in physical health (mean rank=141.14) compared to those with a poor QoL (mean rank=121.49). In contrast, a significant disparity was observed in the psychological domain (Mann-Whitney U test=3135.00, Z= -4.242, p<0.001), suggesting that better psychological well-being was strongly associated with an enhanced QoL. Likewise, social relationships showed a statistically significant difference (Mann-Whitney U test=3996.00, Z= -2.372, p=0.018), suggesting that stronger social ties had a positive impact on QoL. These findings underscore the greater contribution of psychological and social dimensions, compared to physical health, in shaping the quality of life among elderly people (Table 4).

**Table 4:**
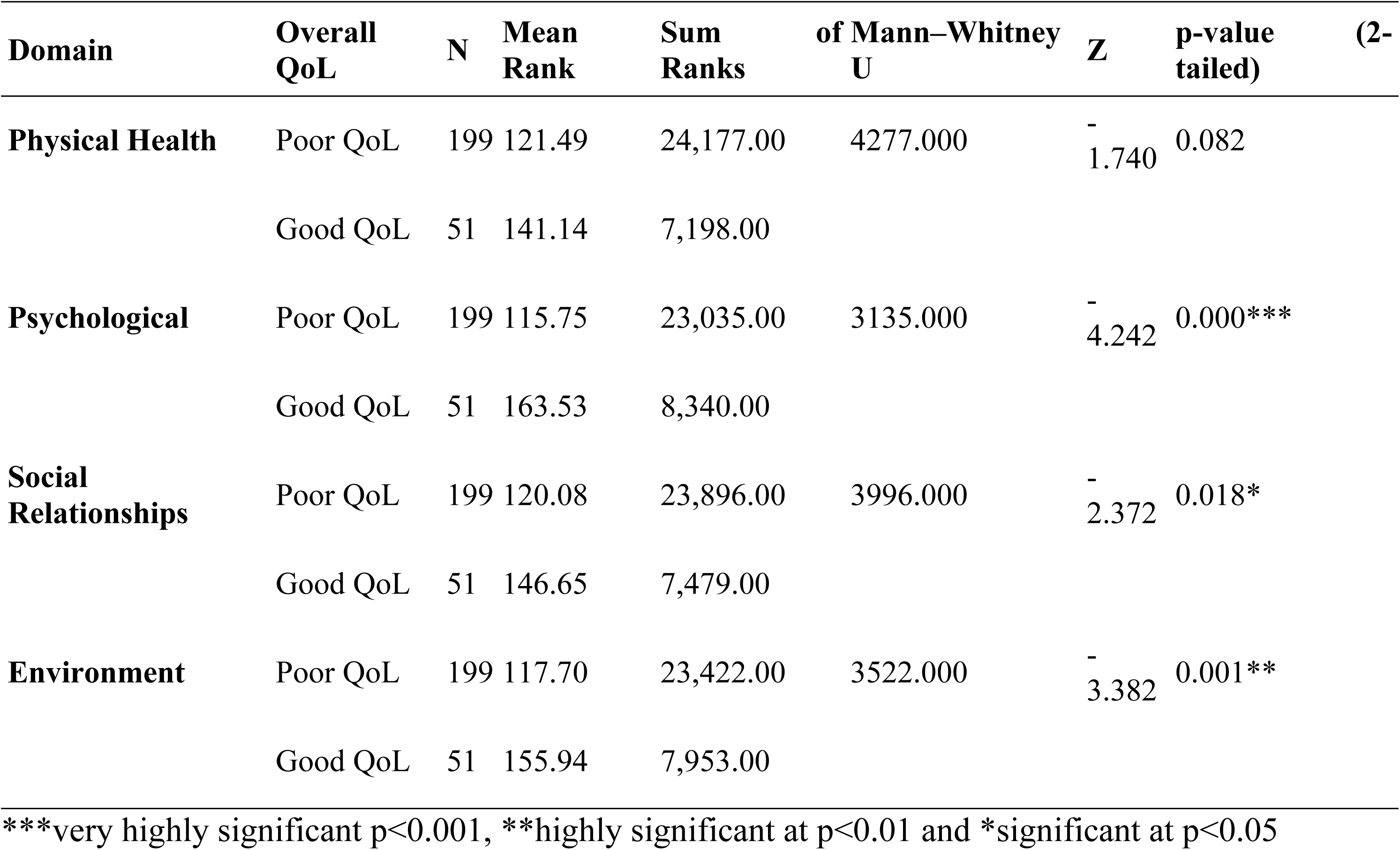
Distribution of Impacts of Physical, Social, Psychological Health on the overall quality of Life (n=250)

The influence of depression on the four different dimensions of QoL was studied (Figure 1). Considering No Depression as a reference category, the Environment dimensions were likely to be impacted 17.37 (7.99-43.70) times for those having a poor quality of Environmental dimensions. Similarly, for Physical QoL, it is likely to be 10.61(5.657-21.38) times for those without depression. Furthermore, the psychological dimension was likely to be affected by 6.30(3.50-11.64) times for those without depression. Moreover, the social dimension was likely to 4.74 (2.53-9.41) times affected those without depression. The impact of depression on Quality of Life was first computed based on the Chi Square test (Annex D).

**Figure 1.**
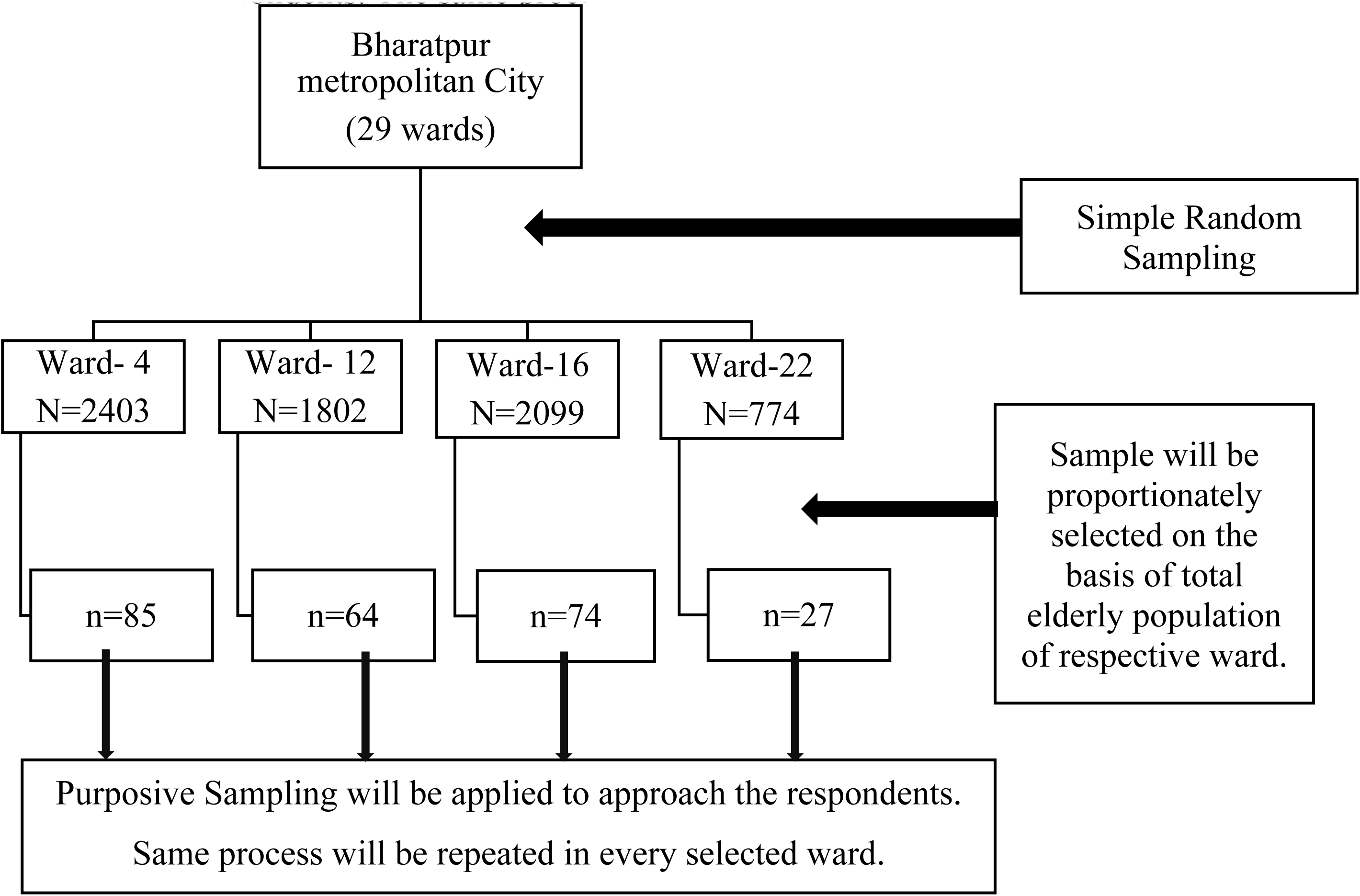
Sampling Procedure for the participation selection in present study

**Figure 2.**
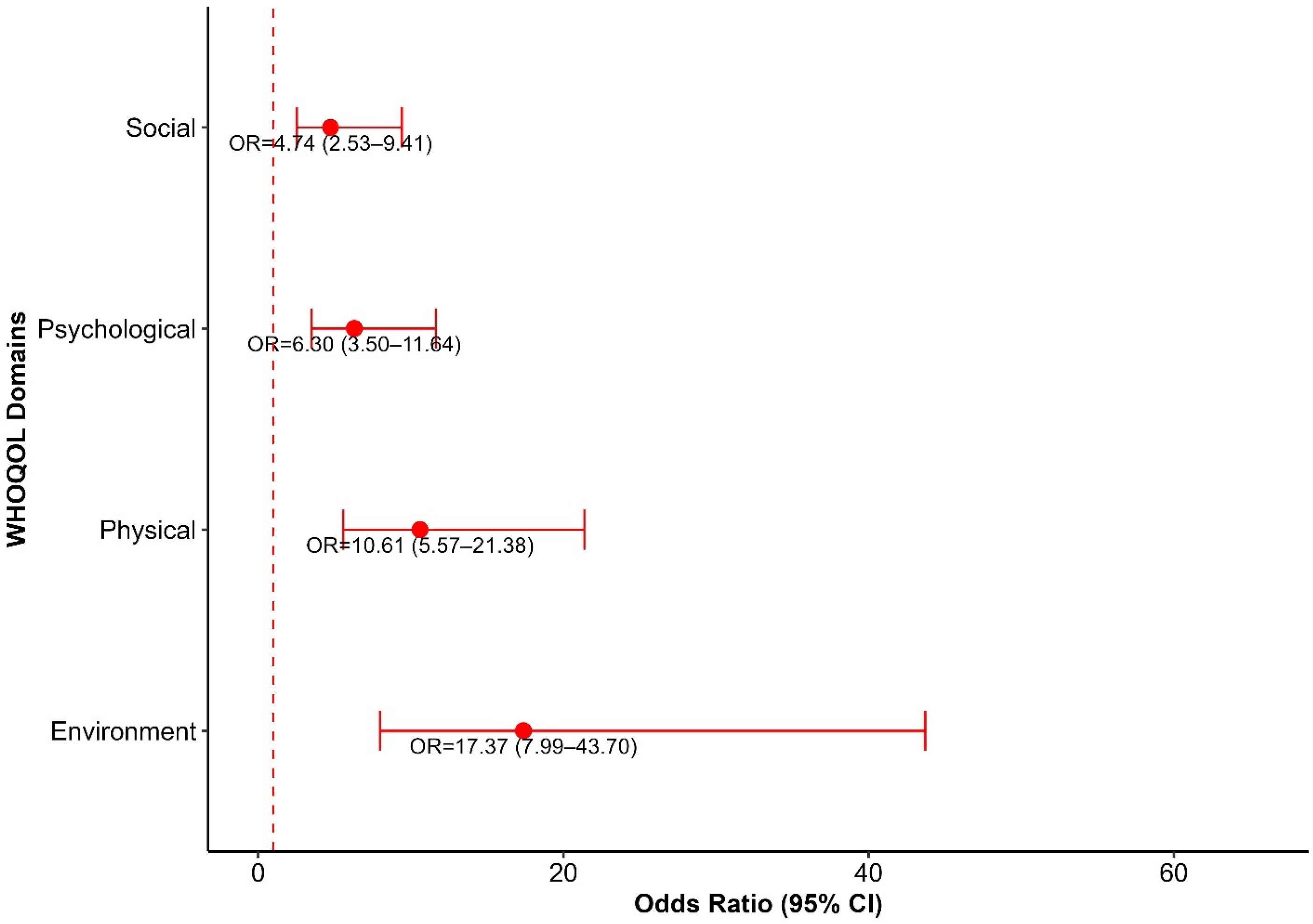
Impact of Depression on Social, Psychological, Physical and Environmental Quality of Life

## Discussion

The primary objective of this study was to explore the status of depression among older adults and to investigate its relationship with various socio-demographic factors in Bharatpur Metropolitan City, situated in the Chitwan District of Nepal. With life expectancy on the rise, the global population is aging, resulting in a higher non-fatal burden of illnesses and injuries, mainly due to the reduction in mortality rates observed from 1990 to 2016.(22) According to our research, depression was substantially linked to overall quality of life (QoL) among the city’s senior citizens. Those scored the Environment dimensions were likely to be impacted seventeen times more for those having a poor quality of the Environmental dimension, for Physical QoL, it is likely to have ten times the psychological dimension was likely to be affected six times for those without depression and the social dimension was likely to more than four times clinically depressive symptoms compared to those without depression even after controlling the comorbidities age, sex and socio-economic level, the negative correlation remained.

These results aligned with many community-based studies conducted in Nepal that show a high incidence of depression and a strong inverse relationship between depression and QoL among senior citizens. Recent research in Nepal, for instance, found that depression was equally common and was linked to Lower QoL. The prevalence of depression was higher in a prior study conducted in rural Nepal (57.4%) than in 28.8% our study. (23) and Kathmandu (24). Furthermore, this study investigated the impact of depression on social, psychological, physical, and environmental QoL, revealing that the environmental dimension is more likely to be affected in those with poor environmental quality. Likewise, individuals without depression were more likely to have better physical QoL.

Our study adds novel insights to the existing literature, which was conducted in various international settings, and shows the prevalence as studies conducted in Japan (25) and Ethiopia (26) shows similar findings to our study. Due to rapid urban growth and shifts in culture within the country, the traditional extended family model has gradually shifted from joint family systems to nuclear family setups.(11) The research found a notable relationship between the education levels of older adults and their experiences of depression. Contrary to our findings, previous research conducted in Nepal has also found a statistically significant link between educational attainment and depression in older adults(27). This study aimed to explore the impact of various physical, social, and mental health factors on overall life quality. The findings revealed that individuals with a higher quality of life exhibited a superior mean rank for physical health compared to those with a lower quality of life. Furthermore, a notable difference was observed in the psychological domain, suggesting that improved mental well-being was closely associated with a better quality of life.

In a similar vein, social connections showed a statistically significant impact, suggesting that stronger social ties positively affect quality of life (QoL). These results highlight the more substantial role of psychological and social factors, as opposed to physical health, in determining the quality of life among the elderly, which aligns with a study conducted in Nepal.(28) Our research aligns with various other studies due to the use of the GDS-15 scale and WHOQOL-BREF, which ensure methodological consistency and produce results consistent with both national and international studies. The cultural context in Nepal, where older adults often face loss and diminished family interaction, may also contribute to similar trends in depression and perceived well-being. This consistency in findings strengthens the global understanding that depression adversely impacts the quality of life in older adults.

Our findings indicate a lower prevalence compared to Pakistan (40.6%) and India (46.2%). This discrepancy in prevalence rates may be attributed to differences in study methodologies, the employment of various depression assessment tools (GDS-5, 15, 30), characteristics of the populations studied, geographical influences, and distinct cut-off points for the GDS score(29).National prevalence remains higher than international figures, potentially due to the pervasive stigma surrounding mental health, insufficient understanding of mental illnesses, limited access to comprehensive mental health services, and the normalization of depression among the elderly, which results in the disease being under-recognized and inadequately treated.

In several cities across Nepal, including Bharatpur, the relationship between depression and quality of life is likely worsened by insufficient mental health resources, societal stigma, and obstacles to accessing care. Studies conducted in various parts of Nepal indicate that factors such as the prevalence of chronic illnesses, limited social support, and the cost and availability of healthcare services play a significant role in this connection. Our research highlights the importance of regular mental health screenings and counseling for the elderly in Bharatpur’s primary care and community environments. Enhancing QoL could be achieved by integrating community-based psychological interventions, like social group activities and physical exercise programs, with elder health checkups, referral pathways, and brief depression screenings, such as the GDS-15.

## Strength and limitations

This research highlights several significant insights. It evaluates both depressive symptoms and quality of life, offering a thorough understanding of the psychological and social dimensions of aging. Conducted among the elderly in community environments, this study delivers practical insights into their mental well-being. The application of standardized tools (GDS-15 and WHOQOL-BREF) ensured the reliability and comparability of the results with other research. These findings have practical applications for community health initiatives, mental health strategies, and elderly care services in Nepal, providing valuable information for healthcare workers, local authorities, and policymakers.

The study was carried out in specific regions of the Bharatpur Metropolitan City with only 250 participants, which may not fully represent the elderly population throughout Nepal. Cultural influences and stigma might have led to the underreporting of psychological distress. Geographic constraints and unmeasured confounding factors further limit the generalizability of our results. Nevertheless, this study establishes a basis for future investigations into the mental health of Nepal’s elderly population.

## Recommendations

Extensive screening programs play a crucial role in detecting unrecognized cases of depression among the elderly. Elderly social groups can encourage the exchange of thoughts and issues, thus offering a reliable social support system. These screening efforts should focus on older adults with chronic illnesses. This research supports improving the physical, mental, and social well-being of seniors while enhancing their overall quality of life. It also suggests training primary healthcare workers to identify depression symptoms and offer suitable counseling or referrals. So, other epidemiological studies should focus on collecting information from larger groups.

## Conclusion

Our study reported the higher status of depression among the elderly population in Bharatpur Metropolitan City, Chitwan, Nepal. The results highlight the considerable influence of psychological and social factors on the quality of life for older adults, rather than their physical health. This research emphasizes the need to improve the overall well-being and health of the elderly. It also points to the necessity of training community-based and primary-level health workers to recognize signs of depression and offer basic counseling or referrals. Additionally, it is crucial to prioritize screening programs for elderly individuals with chronic conditions. This study advocates targeted initiatives to improve the physical, psychological, and social well-being of the elderly.

## Data Availability

All the data used in this analysis are publicly available for further review at https://doi.org/10.7910/DVN/JMHORK

